# Relationship between plasma cortisol concentration and Long COVID symptoms in the post-acute phase of COVID-19: a cross-sectional study and recommendations for future research

**DOI:** 10.1101/2024.11.07.24316777

**Authors:** Thomas Dalhuisen, Halle Grebe, Khamal Anglin, Scott Lu, Sarah A. Goldberg, Lucas Kallás-Silva, Joshua Hauser, Emily Conway, Marin Ewing, Jessica Y. Chen, Emily A. Fehrman, J. Daniel Kelly, Jeffrey N. Martin, Peter W. Hunt, Timothy J. Henrich, Matthew S. Durstenfeld, Steven G. Deeks, Elizabeth Murphy, Morris Schambelan, Michael J. Peluso

## Abstract

**BACKGROUND:** Low cortisol concentrations have been reported in some people with Long COVID (LC), but more data from diverse cohorts are needed to validate this observation. A subset of people with LC present with symptoms resembling those of myalgic encephalomyelitis/chronic fatigue syndrome (ME/CFS). The objective of this study was to compare cortisol concentrations in those with and without Long COVID, with a particular focus on people experiencing ME/CFS-like Long COVID.

**METHODS:** We measured plasma cortisol in 200 individuals 3-6 months following a SARS-CoV-2 infection. Banked biospecimens collected between 8 AM-12 PM were used. Participants met the case definition for Long COVID if they had ≥1 COVID-attributed symptom at least 3 months after symptom onset. People who did not report any symptoms at least 3 months after symptom onset served as recovered controls. Adapting the 2015 Institute of Medicine criteria for ME/CFS, we further defined those with LC resembling ME/CFS (LC-ME).

**RESULTS:** We found no difference in overall morning cortisol concentrations between people with LC (n=144) and those who fully recovered (n=56) (median 8.9 µg/dL vs. 8.8 µg/dL, p=0.97). Analyses of samples collected between 8-10 AM, however, revealed that, compared to those who fully recovered, cortisol concentrations were lower between 8-9 AM for those with LC-ME (median 8.2 vs. 14.8, p=0.02), but higher between 9-10 AM for those with severe LC (>=5 symptoms) (median 12.4 vs. 8.5, p=0.009) and those with LC-ME (median 13.7 vs. 8.5, p=0.02).

**CONCLUSION:** We found no difference in overall morning plasma cortisol concentrations between those with and without Long COVID. Although our data could be suggestive of altered morning cortisol dynamics in a subset of people with Long COVID, longitudinal measures of cortisol in individuals with Long COVID will be critical to further inform the biology of the condition.

## BACKGROUND

Long COVID is a multisystem condition characterized by a wide range of symptoms that can persist for months or years after SARS-CoV-2 infection^1^. The biological mechanisms underlying Long COVID are incompletely understood. Virus persistence, coagulation dysfunction, and inflammation have all been implicated as potential causes^2,3^.

Long COVID is characterized by a variety of different clinical phenotypes and symptom complexes^2,4^. A subset of people with Long COVID experience fatigue, post-exertional malaise, sleep disturbance, neurocognitive dysfunction, and orthostatic intolerance. These symptoms are also characteristic of myalgic encephalomyelitis, sometimes called chronic fatigue syndrome (ME/CFS)^5^. Both Long COVID and ME/CFS are infection-associated chronic conditions (IACCs), which are believed by many to be set off by an infectious trigger^6^. Determining whether post-COVID ME/CFS represents the same condition as ME/CFS that preceded the COVID-19 pandemic (“pre-pandemic ME/CFS”), and whether they share common biology, remains a key question for the field^7^.

SARS-CoV-2 exhibits adrenal tropism, and subsequent adrenal damage and inflammation have been shown in patients with COVID-19^8^. Specifically, SARS-CoV-2 has been found to replicate in the ACE2 and TMPRSS2 positive cells of the adrenal cortex, resulting in reduced zonation, microthrombi formation, and increased inflammatory infiltrates, which consequently could affect cortisol secretion^8^. Differences in systemic cortisol concentrations have been identified in people with Long COVID, suggesting a dysfunction of the hypothalamic-pituitary-adrenal (HPA) axis^9^ in this condition. One cross-sectional study, for example, revealed significantly lower cortisol in people with Long COVID compared to healthy controls^9^. In fact, cortisol was the single strongest predictor of whether an individual in that study had Long COVID. Other studies, however, have found no evidence for lower cortisol in people with Long COVID^10,11^. Interestingly, studies of pre-pandemic ME/CFS reveal similarly inconsistent results regarding cortisol, with some providing evidence for lower cortisol concentrations and others not^12,13,14^. A caveat for the interpretation of all of these studies is that cortisol is secreted in a pulsatile manner and there is significant diurnal variation with the highest concentrations in the morning and lowest at night. Thus, the timing of collection may play an important role in the interpretation of results. In addition, the secretion of endogenous cortisol can be modified by exposure to exogenous glucocorticoids, which are commonly used among the population in general and among some individuals with Long COVID.

To further investigate these questions, we conducted a pilot study to cross-sectionally measure morning plasma cortisol concentrations in a cohort of 200 highly characterized individuals 3-6 months post-COVID. Our objective was to compare cortisol concentrations in those with and without Long COVID with a particular focus on people experiencing ME/CFS-like Long COVID.

## METHODS

### Study Design

In this cross-sectional study, we measured plasma cortisol concentrations in participants 3 to 6 months following symptom onset from a SARS-CoV-2 infection. We included participants who had their blood collected between 8 AM and 12 PM.

### Participants

Participants were volunteers in the University of California, San Francisco (UCSF)-based Long-term Impact of Infection with Novel Coronavirus (LIINC) research program (NCT04362150)^15^. Study procedures have been described in detail previously^15^. LIINC enrolls participants with test-confirmed SARS-CoV-2 infection at least two weeks following their symptom onset date. Participants are examined and complete an interviewer-administered symptom questionnaire at an initial study visit and approximately every four months thereafter.

### Case definitions

For these analyses, we defined three groups: a non-Long COVID (nLC) group, a Long COVID (LC) group, and a ME/CFS-like Long COVID (LC-ME) group. The nLC group consisted of participants who did not report any COVID-attributed symptoms in the post-acute phase and were considered to have fully recovered from their infection. We defined Long COVID according to the World Health Organization consensus criteria^16^, as one or more COVID-attributed symptom present for more than 3 months following their initial SARS-CoV-2 symptom onset date. We further defined those with Long COVID who met an adaptive version of the 2015 Institute of Medicine ME/CFS criteria^5^ as having LC-ME. These participants reported fatigue, sleep disturbance and neurocognitive dysfunction or orthostatic intolerance. Of note, at the time of this pilot study, a dedicated question regarding post-exertional malaise had not yet been added to the study questionnaire.

### Measurements

Interviewer-administered questionnaires were used to collect data on Long COVID symptomatology; importantly, symptoms that pre-dated the COVID-19 diagnosis and were unchanged were not recorded as representing Long COVID. Cortisol concentrations (μg/dL) were measured using previously banked blood samples using the Siemens Atellica® IM Analyzer. Cortisol was measured at the UCSF Zuckerberg San Francisco General Hospital Clinical Laboratory.

### Statistical Analyses

Cortisol concentrations between groups were compared using Wilcoxon rank-sum test. We considered a p-value of <0.05 as statistically significant. All analyses were carried out with R software version 4.2.2^17^.

## RESULTS

### Study participants

We measured cortisol in 200 participants 3 to 6 months following a SARS-CoV-2 infection. Symptom onset date ranged from March 2020 to October 2022. 144 (72%) participants met our case definition for Long COVID at the post-acute timepoint. In comparison to the nLC group, the Long COVID group had a similar age (median age 48 vs. 44 years, p=0.5), distribution of birth sex (50% vs. 56%, p=0.4) and percentage who had been vaccinated prior to SARS-CoV-2 infection (48 % vs. 51%, p=0.9). A small number of Long COVID participants (3%) reported exogenous glucocorticoid use. Median blood collection time for cortisol measurements was similar for both groups (10:18 AM vs. 10:30 AM). Participant characteristics are shown in **Table 1**.

**Table 1.**
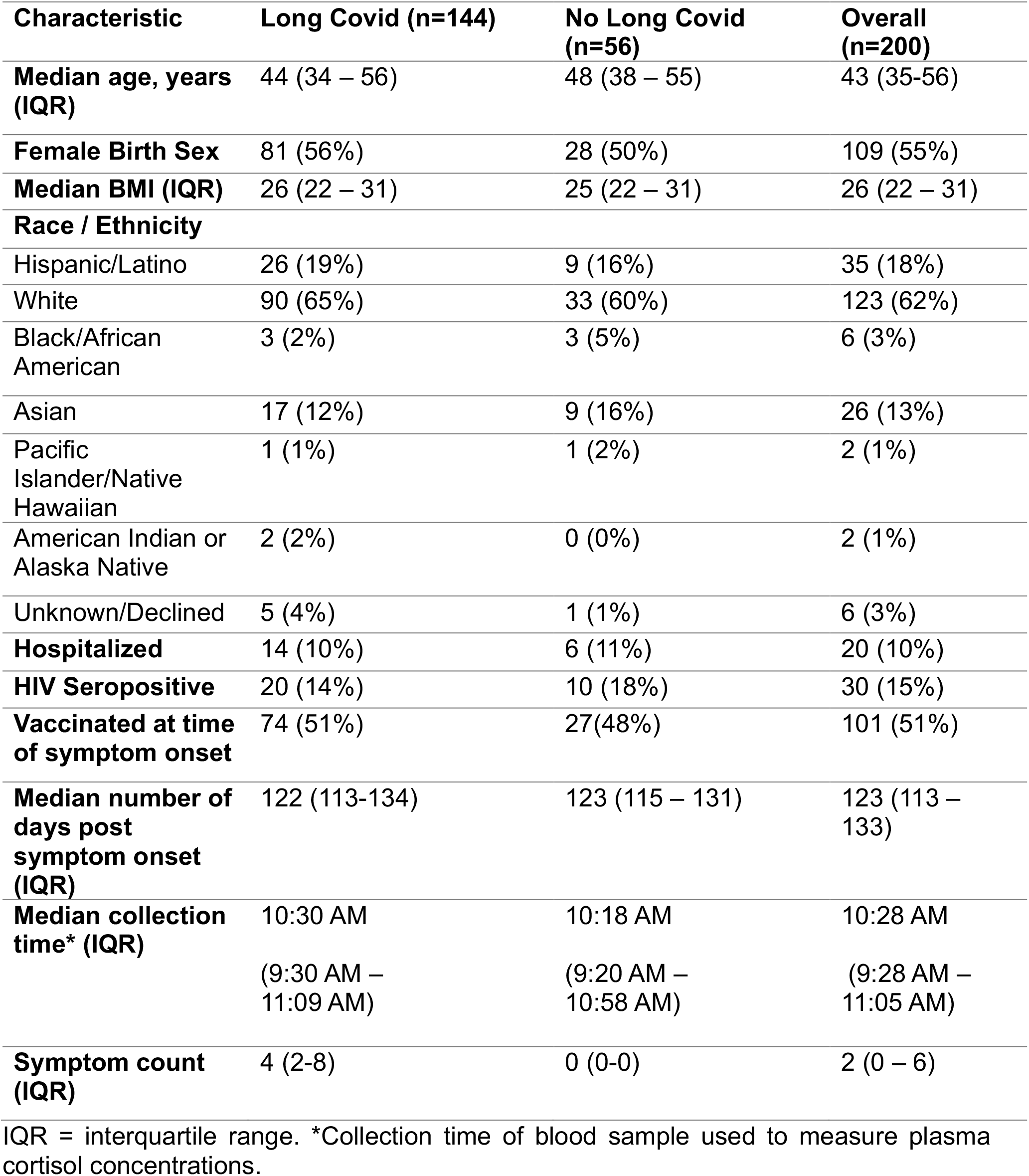
Characteristics of study participants.

| **Characteristic** | **Long Covid (n=144)** | **No Long Covid (n=56)** | **Overall (n=200)** |
| --- | --- | --- | --- |
| **Median age, years (IQR)** | 44 (34 – 56) | 48 (38 – 55) | 43 (35-56) |
| **Female Birth Sex** | 81 (56%) | 28 (50%) | 109 (55%) |
| **Median BMI (IQR)** | 26 (22 – 31) | 25 (22 – 31) | 26 (22 – 31) |
| **Race / Ethnicity** |  |  |  |
| Hispanic/Latino | 26 (19%) | 9 (16%) | 35 (18%) |
| White | 90 (65%) | 33 (60%) | 123 (62%) |
| Black/African American | 3 (2%) | 3 (5%) | 6 (3%) |
| Asian | 17 (12%) | 9 (16%) | 26 (13%) |
| Pacific Islander/Native Hawaiian | 1 (1%) | 1 (2%) | 2 (1%) |
| American Indian or Alaska Native | 2 (2%) | 0 (0%) | 2 (1%) |
| Unknown/Declined | 5 (4%) | 1 (1%) | 6 (3%) |
| **Hospitalized** | 14 (10%) | 6 (11%) | 20 (10%) |
| **HIV Seropositive** | 20 (14%) | 10 (18%) | 30 (15%) |
| **Vaccinated at time of symptom onset** | 74 (51%) | 27(48%) | 101 (51%) |
| **Median number of days post symptom onset (IQR)** | 122 (113-134) | 123 (115 – 131) | 123 (113 – 133) |
| **Median collection time\* (IQR)** | 10:30 AM<br>(9:30 AM – 11:09 AM) | 10:18 AM<br>(9:20 AM – 10:58 AM) | 10:28 AM<br>(9:28 AM – 11:05 AM) |
| **Symptom count (IQR)** | 4 (2-8) | 0 (0-0) | 2 (0 – 6) |

IQR = interquartile range. \*Collection time of blood sample used to measure plasma cortisol concentrations.

### Cross-sectional analysis of morning cortisol concentrations

We first compared morning plasma cortisol concentration (µg/dL) aggregated across all sample collection time points between 8 AM and 12 PM (**Fig. 1A**). We found no difference in median morning cortisol between the LC and nLC group (median 8.9 vs. 8.8, p=0.97). Cortisol concentrations below the lower limit of normal (<5 µg/dL) were found in 14 (9.7%) LC participants and 5 (8.9%) nLC participants.

**Figure 1.**
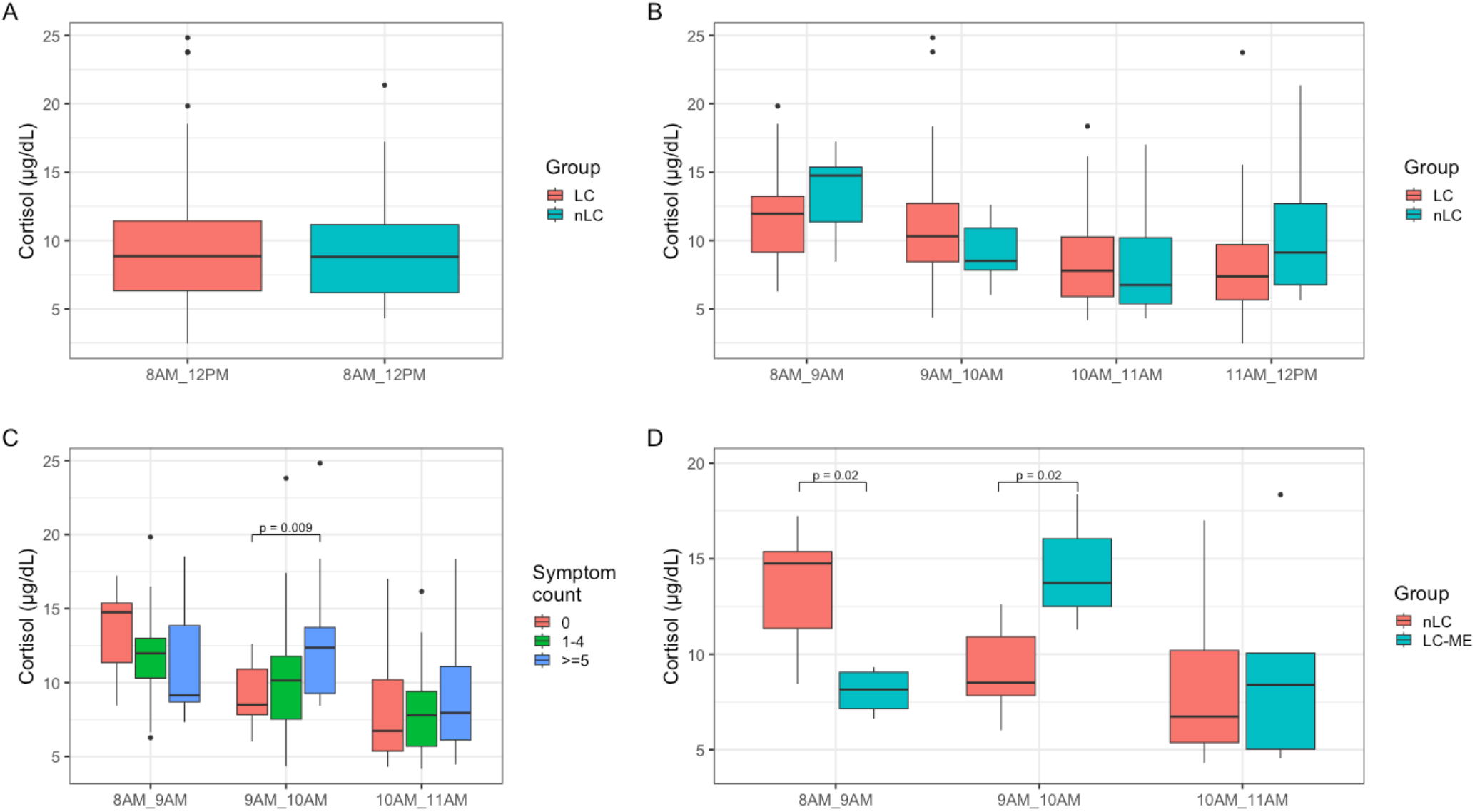
A. Morning cortisol concentration (µg/dL) in participants with Long COVID compared to those without. **B**. Cortisol concentrations stratified by hourly time bin. **C**. Cortisol concentrations stratified by symptom count and hourly time bin. Statistically significant group comparisons are indicated. **D**. Cortisol concentrations in LC-ME compared to those without stratified by hourly time bin. Statistically significant group comparisons are indicated. Abbreviations: LC = Long COVID; nLC = non-Long COVID (e.g., post-COVID but fully recovered); LC-ME = ME/CFS-like Long COVID according to the adapted 2015 Institute of Medicine criteria.

### Cross-sectional analysis of cortisol concentrations by hour

Given the dynamic nature of cortisol secretion, we next examined cortisol concentrations stratified by hour of collection (**Fig. 1B**). We observed a non-significant trend for lower cortisol concentrations in those with LC between 8-9 AM (n=19), compared those without LC (n=7) (median 12.4 vs.14.8, p=0.36). Between 9-10 AM, however, this relationship reversed, and cortisol concentration tended to be higher in the LC group (n=33) compared to the nLC group (n=12) (median 10.3 vs. 8.5, p=0.15), though again these differences were not significant. Cortisol concentrations between 10-11 AM and 11 AM-12 PM were similar in both groups.

### Cortisol concentrations by symptom count

We next stratified the LC group into a severe and non-severe group. Participants in the severe LC group reported 5 or more symptoms (n=60), while participants in the non-severe LC reported between 1-4 symptoms (n=84) (**Fig. 1C**). The severe LC group had a non-significant trend towards lower cortisol concentration between 8-9 AM (n=9) (median 9.1) compared to the non-severe LC group (n=13) (median 12.0, p=0.52) and the nLC group (n=7) (median 14.8, p=0.44) However, between 9-10 AM, the severe LC group had the highest cortisol concentrations, which differed significantly from the nLC group (median 12.4 vs. 8.5, p=0.009). Cortisol levels were comparable between all groups after 10 AM.

### Cortisol concentrations in participants with ME/CFS-like Long COVID

Finally, we conducted an exploratory analysis to determine whether the observed differences were driven by the ME/CFS-like phenotype of Long COVID (**Fig. 1D**). Cortisol concentrations between 8-9 AM were significantly lower in the LC-ME group (n=7) compared to the nLC group (n=12) (median 8.2 vs. 14.8, p=0.02). Conversely, cortisol concentrations between 9-10 AM were significantly higher in the LC-ME group (n=3) compared to the nLC group (n=12) (median 13.7 vs. 8.5, p=0.02). This difference was not observed when comparing the nLC group with those with LC but without ME/CFS (14.8 vs. 12.6, p=0.78, 8-9 AM; 8.5 vs. 10.2, p=0.27, 9-10 AM). Cortisol levels were comparable between all groups after 10 AM.

## DISCUSSION

Although we were unable to confirm the broad group differences in morning plasma cortisol concentrations between those with and without Long COVID observed in a prior report^9^, we did make several potentially interesting observations that will inform future work. We found that compared to people who had fully recovered from COVID-19, those with Long COVID tended to exhibit cortisol concentrations that were lower between 8-9 AM but higher between 9-10 AM. This pattern appears to be driven by a more symptomatic subset of participants, including those with ME/CFS-like symptoms. Taken together, our data are suggestive of altered morning cortisol dynamics in a subset of people with Long COVID. Although the etiology and clinical significance of these findings remains unknown, it suggests a potential physiologic pattern that may distinguish one phenotype of Long COVID and could provide clues as to the pathophysiology driving this debilitating condition.

In contrast to a prior report^9^, we failed to detect a difference in cortisol concentration in people with Long COVID. This may be explained by differences in cohort characteristics. For example, the median time since acute infection was substantially longer in the prior study than in our cohort (14 months versus 4 months). It could be that this physiologic difference in cortisol concentrations becomes more pronounced over time; this would be consistent with observations in which the disease state and biomarker profiles of ME/CFS, another infection-associated chronic condition, evolves over time^12^. Another more likely possibility, however, is related to heterogeneity in sampling collection time. Cortisol concentrations fluctuate during the day, typically peaking early in the morning after waking followed by a rapid decline^18^. It is possible that inconsistencies in sample timing, even if relatively minor, could drive observable differences if one group is measured later than another. In fact the prior report did note differences in sampling time between Long COVID and control groups^9^, although the differences did not reach statistical significance.

Our observation of what may be a different pattern of cortisol secretion in those with ME/CFS-like Long COVID is suggestive of a delayed morning cortisol peak. Although participants in our LC-ME group were not formally diagnosed with ME/CFS, similar examples of alterations in cortisol dynamics can be found the ME/CFS literature^12,13,19^. For example, one early study measured longitudinal blood cortisol at multiple timepoints throughout the day in patients with ME/CFS and found a reduction in early-morning cortisol^13^. A more recent study found significantly lower salivary cortisol secretion in the morning and flattened cortisol secretion curves at 4 collection times throughout the day among ME/CFS patients compared to healthy controls^20^. Overall, our findings suggest that further investigation of the patterns of cortisol secretion among those with Long COVID, particularly of the ME/CFS-like phenotype, are warranted.

This study has several limitations. We did not collect information on the time of awakening, therefore analyses could not adjust for waking time. Therefore, these results could be explained by those with ME/CFS having a later waking time and therefore later peak. Our applied case definition of ME/CFS, while consistent with the 2015 IOM criteria, did not accurately capture post-exertional malaise symptoms because this symptom was inconsistently recorded in the first few years of our study; this may have resulted in misclassification of individuals as having ME/CFS-like Long COVID, although such misclassification would be expected to make it more difficult to observe an effect. Furthermore, our follow-up interval (3-4 months) represents an earlier timescale than is required for a formal diagnosis of ME/CFS (6 months). We also acknowledge small sample sizes when comparing cortisol concentrations cross-sectionally within the group strata in this exploratory analysis.

While we believe these observations are intriguing, the main limitation of our study is that these cross-sectional findings do not capture the secretory pattern in any one patient. To better understand cortisol dynamics in Long COVID, future studies should focus on longitudinal measurements of cortisol within individuals. For example, performing serum or salivary collection at short intervals over the course of a day would provide a better assessment of the overall secretory patterns. It is also important that future studies take into consideration the many factors that can have a direct impact on HPA axis activity and cortisol secretion, including glucocorticoid use which is particularly prevalent among those with this disease condition. It will be vital to take these factors into account to improve study validity and generalizability of results. **Table 2** outlines some considerations we deem important for future studies.

**Table 2.** Proposed approaches or considerations for studies evaluating cortisol secretion dynamics.

| **Confounding Variable** | **Proposed Approach or Considerations** |
| --- | --- |
| **Medications** | Obtain a full medication list and carefully document participants' medications, especially those which may alter cortisol secretion (e.g., glucocorticoids, levothyroxine, etc.)<sup>21</sup>. Obtain full medication list, as many participants may not be aware that a medication they are taking is a glucocorticoid. Consider exclusions based on current or recent use. |
| **Phase in menstrual cycle** | If applicable, record the participant's menstrual cycle phase. Some data suggest cortisol awakening response levels increase during ovulation<sup>22</sup> and an increased glucocorticoid<sup>23</sup> response in the luteal phase. |
| **Sleep cycle disturbances** | Record participants' usual sleep patterns. Abnormal sleep patterns, like those often associated with shift work, alter the body's circadian rhythms and may affect the timing of cortisol secretion<sup>24</sup>. |
| **Mental health** | Carefully document participants' mental health comorbidities. Individuals with depression have been shown to have a hyperactive HPA axis<sup>25</sup>. |
| **Intercurrent infections** | Screen for and document any recent or intercurrent infections. Cortisol secretion may be altered in chronic<sup>26</sup> and acute infections, including COVID-19<sup>27</sup>. |
| **Substance use history** | Document recent history of substance use. Alcohol, nicotine and other drugs have been implicated in dysregulation of cortisol secretion<sup>28,29,30</sup>. |
| **Recent travel across time zones** | Exclude participants who have recently traveled across time zones<sup>31</sup>. |
| **Comorbidities** | Document history of liver or kidney disease, which may directly impact cortisol secretion and/or metabolism<sup>32,33,34,35</sup>. |
| **Abnormal cortisol binding globulin (CBG)** | Consider testing samples for differences in quantity of CBG, which may vary significantly between individuals<sup>36</sup>. |

Overall, while we could not confirm the dramatic differences in cortisol values observed in prior work, we observed differences in cortisol secretory patterns that suggests that continued efforts to understand pathways related to the HPA axis in Long COVID may be informative. Future efforts will be crucial in eliciting the role of altered cortisol dynamics in individuals with Long COVID and will continue to inform the pathophysiology associated with this condition.

## Data Availability

All data produced in the present study are available upon reasonable request to the authors.

## Acknowledgements

We are grateful to the study participants and their medical providers. We acknowledge current LIINC clinical study team members Brent Abel, Grace Anderson, Kofi Asare, Tyrine Bailey, Melissa Buitrago, Celina Chang Song, Alexus Clark, Avery Eun, Tony Figueroa, Diana Flores, Yuki Fudotan, Rebecca Hoh, Beatrice Huang, Megan Lew, Michael Luna, Kathleen Bellon Pizzaro, Douglas Robbins, Antonio Rodriguez, Dylan Ryder, Viva Tai, Julian Uy, Meghann Williams, and Andhy Zamora; and current LIINC laboratory team members Belen Altamirano-Poblano, Amanda Buck, Lilian Grimbert, and Brian LaFranchi. We thank Pahul Chhabra, Aidan Donovan, Carrie Forman, Rania Ibrahim, Marine Lyden, Sean Thomas, and Badri Viswanathan for assistance with data entry and review. We also acknowledge all former LIINC clinical, laboratory, and data team members. We thank the UCSF AIDS Specimen Bank for processing specimens and maintaining the LIINC biospecimen repository. We are grateful to Elnaz Eilkhani and Monika Deswal for regulatory support. We are also grateful for the contributions of additional LIINC leadership team members: Matthew Durstenfeld, Priscilla Hsue, Bryan Greenhouse, Ma Somsouk, Isabelle Rodriguez-Barraquer, and Rachel Rutishauser.

## FOOTNOTES

### Funding

MJP is supported on K23AI157875. The LIINC clinical core is supported by the PolyBio Research Foundation, with additional funding from R01AI141003 (to TJH) and 1R01NS136197 (to MJP).

### Conflicts of interest

SGD reports consulting for Enanta Pharmaceuticals and Pfizer and reports research support from Aerium Therapeutics outside the submitted work. MJP has received consulting fees from Gilead Sciences, AstraZeneca, BioVie, Apellis Pharmaceuticals, and BioNTech and research support from Aerium Therapeutics and Shionogi, outside the submitted work.

## Notes

### Author Declarations

This study was approved by the UCSF Institutional Review Board.

